# Characteristics, treatment and outcomes of myelodysplastic syndrome in two Finnish hospital districts

**DOI:** 10.1101/2023.10.06.23296265

**Authors:** Samuli Tuominen, Tatu Miettinen, Christina Dünweber

## Abstract

**Introduction:** Myelodysplastic syndromes (MDS) are hematologic malignancies characterized by changes in haematopoiesis and a high risk for progressing into acute myeloid leukemia (AML). In this retrospective registry based real-world study, from two Finnish hospital data lakes we characterized specialised health care treated MDS patients, their treatment landscape, outcomes, and healthcare resource utilization.

**Methods:** This study consisted of adult patients with MDS diagnosed in either of two hospital districts in Finland: hospital district of Southwest Finland (HDSF) and Pirkanmaa hospital district (PHD). Two different time windows were used depending on data availability: 1.1.2010-31.12.2019 (HDSF) and 1.1.2012-31.12.2019 (PHD). Electronic health record data, including demographics, diagnoses, and medications was accessed via the respective hospital data lakes and dates and causes of death data was collected from Statistics Finland.

**Results:** We identified 565 adult MDS patients, of whom 424 received active life-prolonging treatment at specialized healthcare and 141 were treated with watchful observation or supportive care at primary care. 72 patients were treated with azacitidine and 26 patients received allogeneic hematologic stem cell transplant. Median overall survival for the specialty healthcare treated patients was 27,5 months (95 confidence interval [CI] 24,1-35,2) and costs per patient year were 17 563€.

**Conclusion:** This hospital data lake-based analysis identified patient groups with differing disease severity and need for treatment. High-risk, azacitidine treated patients have suboptimal outcomes and high costs, highlighting the need for new therapeutic approaches to prevent disease progression and reduce disease burden.

## Introduction

Myelodysplastic syndromes (MDS) are a heterogenous group of clonal haematological malignancies where the production of normal blood cells is defective, leading to both a deficiency of mature hematopoietic cells, as well as increased blast cells. MDS is clinically manifested as cytopenias, and a risk to develop acute myeloid leukemia (AML). Patients, often older adults, suffer from anaemia and infections and often become dependent on red blood cell (RBC) transfusions.

Treatment of MDS patients consists of few options. The only chance for curative treatment of MDS is allogeneic hematopoietic stem cell transplant (HSCT), but unfortunately most patients are not fit for this procedure (1). For low risk non-symptomatic patients watchful observation is used. For high-risk patients unable to undergo HSCT, the hypomethylating agent azacitidine is used. Lenalidomide is used for patients positive for Del(5q). Fragile patients only receive supportive care including RBC transfusions and erythropoietins.

Exciting real-world studies on MDS have described survival and HCRU before (2–4). However, the Finnish hospital data lakes and the ability to link detailed hospital patient data with other registries (here causes of death data from Statistics Finland) provide a unique aspect into an integrated understanding of the patients, their treatments, comorbidities and prognosis.

As the diagnosis of MDS is attempted earlier in the disease process, and for all patients, it is important to understand the patient journey to support decisions on which patients to actively and / or aggressively treat, and who benefit most from watchful observation. This study was designed to reveal the treatment pathways of MDS patients and differences in their outcomes and HCRU.

## Methods

### Data access

This was a retrospective study of patients diagnosed with MDS or Chronic Myelomonocytic Leukemia (CMML) in two hospital districts: HDSF and PHD in Finland from January 1^st^ 2010 to December 31^st^ 2019. Findata, the social and health data authority in Finland, approved this study (data permit number THL/2767/14.02.00/2020). According to the Finnish Act on Secondary use of Health and Social data (552/2019) no patient consent was required.

### Patient inclusion criteria/Cohort formation

Patients were included if they were at least 18 years of age at the date of the first recorded MDS or CMML diagnosis code and had a diagnosis code for MDS (D46.*) or CMML (C93.1) recorded during 1.1.2010-31.12.2019 (HDSF) or 1.1.2012-31.12.2019 (PHD). Patients recorded with MDS or CMML prior to the beginning of the inclusion period of the corresponding hospital district were excluded. At PHD diagnoses were available starting from 2010, thus the inclusion period at PHD started at 2012 in order to have 2-year washout period to prevent the inclusion of non-incident patients. In order to prevent inclusion of patients misdiagnosed with MDS during the diagnosis process whose diagnosis was corrected later to another haematological condition, we decided on the following criteria: 1) The diagnosis code had to be marked at least three times at the electronic health records within 6 months of the first diagnosis, and 2) if the patient received a diagnosis code for D47 (other neoplasms of uncertain behaviour of lymphoid, hematopoietic and related tissue), C90 (multiple myeloma), C92.1 (chronic myeloid leukemia), C81-C88 (lymphomas) or D45 (polycythemia vera) and MDS was no longer recorded after that, the patient was excluded 3) if the patient died within 6 months of first MDS diagnosis, they were included regardless of criteria 1) and 2). The date of the first record of the MDS or CMML diagnosis code was defined as the index. The length of follow-up was defined as time from index until death, progression to AML (first record of C92.0), or end of study (31.12.2019).

### Subgroup formation

Patient age and sex along with all ICD-10 diagnosis codes registered during the health care contacts, medical treatments with time stamp, generic names and ATC-codes, procedure codes and records of laboratory measurements were obtained from the HDSF and PHD data lakes. Patients were stratified into 2 subgroups: 1) patients treated at the specialty health care (SHC) and 2) patients who were untreated and only received supportive care or who were only diagnosed at SHC and possibly treated at primary health are (PHC). As the PHC data was not available for this study, further assessing the latter patient group was not feasible. Two mutually distinct subgroups were distinguished from the patients treated at SHC, namely patients who were detected receiving allogeneic haemotopoietic stem cell transplant (HSCT) and patients receiving azacitidine treatment. Note that some of the patients in the HSCT subgroups also received azacitidine, however, this was used either as an induction therapy/treatment for the transplant or as maintenance/relapse treatment after the transplant. Only transplants and azacitidine treatments prior to potential progression to AML were considered. In order to detect the stem cell transplants, three criteria were applied: 1) procedure recorded at the data lake (WW302, WW304 or WW306), 2) text specifier recorded alongside the Z94.8 diagnosis code (“Other transplanted organ and tissue status”) mentioning the exact date of the transplant, or 3) immunosuppressive medication combined with a prior HLA-testing. Immunosuppressive medications included were: ciclosporin, everolimus, tacrolimus, mycophenolic acid and methotrexate. Date of the first record of the immunosuppressive medication was used as the date of the transplant as the immunosuppressive treatment is typically initiated day before or the day of transplant.

### Patient characteristics & treatments

Number and proportion of patients per sex and age group in 10-year intervals were reported. Number and proportion of patients with either a malignancy or chemotherapy before MDS or CMML diagnosis were reported. For a previous malignancy it was required that a patient had at least three records of the same cancer diagnosis code (at two-character level i.e., ‘C’ followed by two numbers) on separate days. A previous chemotherapy was defined as any medical treatment with an ATC-code starting with ‘L01’ (excluding hydroxycarbamide; ATC-code L01XX05) at least 3 months before the index. Charlson comorbidity index was calculated for each patient from the ICD-10 diagnosis codes recorded prior to index (5). The number and proportion of patients with CCI 0, 1-2, 3-4 and 5+ were reported.

### Mortality and time to event analyses

Dates of death were obtained from the HDSF and PHD data lakes and causes of death were obtained from the causes of Death (Statistics Finland) register up to the end of study period. Overall survival was defined as time from index until death (event) or end of study (31.12.2019; censoring event). MDS or CMML mortality was defined as any death with MDS or CMML recorded as immediate or main cause of death. Other mortality was defined as death without records of MDS or CMML in either main or immediate causes of death. Date of progression to AML was defined as the first day patient being recorded with C92.0. Overall survival was estimated via Kaplan-Meier fits while progression rate to AML, MDS or CMML mortality and other mortality were assessed simultaneously via Aalen-Johannsen fits, which is an extension of the Kaplan-Meier method taking into account the likely competing risks setting between multiple, alternative outcomes. Event free survival (EFS) was defined as time from index until composite event of either progression to AML or death (any cause).

### Health care resource utilization (HCRU)

All hospitalizations and outpatient contacts of the patients at the specialty health care of HDSF and PHD were obtained. Outpatient contacts per patient year (PPY) and continuous hospitalizations, as well as total number of inpatient days (i.e., days spent at the ward) PPY were calculated. Furthermore, prices of the contacts were joined based on the specialty of the contact. Prices were obtained from Mäklin et al (6). These prices were then used to calculate costs PPY. Here ‘patient’ years correspond to the length of follow-up. Contacts recorded with MDS or CMML diagnosis were defined as MDS or CMML specific HCRU. Confidence intervals were obtained using bootstrapping with 10 000 bootstrap samples.

### Statistical analysis

All data were delivered to and analysed in the Findata secure data analysis environment. Only aggregated results files not containing any patient-level data were pulled from the analysis environment for further reporting. All statistical analyses were run using R: A language and software for statistical computing, version 4.0.3. Time to event analyses were run using the survival R-package, A Package for Survival Analysis in R (7).

## Results

### Patient characteristics

In total 565 patients fulfilling the inclusion criteria, 325 from HDSF and 232 from PHD. As in Finland the diagnosis of MDS is always made in the specialised healthcare setting, this likely represents all MDS patients from these two districts. The median age of the patients was 77 years (interquartile ratio (IQR): 68-83 years), included 223 females (39.5%) and had a median follow-up time of 17 months (IQR: 6-39 months) (Table 1). Incidence of MDS in our cohort was 6,06 per 100 000 persons. Our cohort included 13 patients with diagnosis specifically for CMML, but as the number of these patients was so small, they were not analysed separately. 52 patients (9%) had a previous cancer diagnosis, most prominently prostate cancer (2,8%), breast cancer (1,1%) and other haematological malignancies. 70 (12,4%) patients had previous cancer diagnosis or chemotherapy, indicating secondary or treatment related MDS. Other frequent comorbidities (% during the whole follow-up) included essential hypertension (37,9%), atrial fibrillation (25,3%), heart failure (20,7%) and chronic ischaemic heart disease (19,3%). Charlson comorbidity index (CCI) which classifies and summarises prognostic diagnoses into an integer score (8) where scores of 0-1 are considered low comorbidity burden, and 2 or more is considered increasingly higher. In our data, 36% of patients has CCI score >2.

**Table 1.** Characteristics of MDS patients.

| Variable | Level | All |  | Treated at SHC |  | Not treated at SHC |  | AZA treated |  | HSCT |  |
| --- | --- | --- | --- | --- | --- | --- | --- | --- | --- | --- | --- |
|  |  | N | % | N | % | N | % | N | % | N | % |
| N | - | 565 | 100 | 424 | 75 | 141 | 25 | 72 | 13 | 26 | 5 |
| Age group (at index) | Less than 55 | 28 | 5 | 18 | 4 | 10 | 7 | < 5 | - | < 5 | - |
|  | 55 – 59 | 23 | 4 | 17 | 4 | 6 | 4 | < 5 | - | < 5 | - |
|  | 60 – 64 | 50 | 9 | 39 | 9 | 11 | 8 | 6 | 8 | 13 | 50 |
|  | 65 – 69 | 56 | 10 | 48 | 11 | 8 | 6 | 11 | 15 | 8 | 31 |
|  | 70 – 74 | 76 | 14 | 58 | 14 | 18 | 13 | 22 | 31 | 0 | 0 |
|  | 75 – 79 | 116 | 21 | 83 | 20 | 33 | 23 | 16 | 22 | 0 | 0 |
|  | 80 – 84 | 127 | 23 | 93 | 22 | 34 | 24 | 9 | 13 | 0 | 0 |
|  | 85 + | 89 | 16 | 68 | 16 | 21 | 15 | 0 | 0 | 0 | 0 |
| Sex | Female | 223 | 40 | 169 | 40 | 54 | 38 | 22 | 31 | 9 | 35 |
|  | Male | 342 | 61 | 255 | 60 | 87 | 62 | 50 | 69 | 17 | 65 |
| Charlsson Comorbidity Index (CCI; at index) | 0 | 363 | 64 | 260 | 61 | 103 | 73 | 59 | 82 | 22 | 85 |
|  | 1-2 | 148 | 26 | 123 | 29 | 25 | 18 | 11 | 15 | < 5 | - |
|  | 3-4 | 38 | 7 | 29 | 7 | 9 | 6 | < 5 | - | 0 | 0 |
|  | >=5 | 16 | 3 | 12 | 3 | < 5 | - | 0 | 0 | 0 | 0 |
| Other malignancy or previous chemotherapy | No previous cancer or chemotherapy | 495 | 88 | 367 | 87 | 128 | 91 | 63 | 88 | 24 | 92 |
|  | Previous cancer or chemotherapy | 70 | 12 | 57 | 13 | 13 | 9 | 9 | 13 | < 5 | - |

Treated at SHC=patients receiving active treatment and specialised healthcare, Not treated at SHC= patients who do not receive active treatment at specialised healthcare, AZA treated=azacitidine treated patients, HSCT= HSCT patients who received HSCT.

141 patients (25%) initially diagnosed with MDS at specialised healthcare (SHC), received no further treatment at specialised healthcare (Table 1., “Not treated at SHC”). Most of these patients likely received treatment at primary care setting, of which we have no data. These patients include low risk, non-symptomatic patients who are usually treated with watchful observation or receive supportive care.

### Treatment of MDS patients at specialty health care

Only a handful of medications are used to treat MDS in Europe and worldwide (9). HSCT is the only potentially curative treatment for MDS patients, but many MDS patients are not suitable candidates for it, due to old age, comorbidities, poor performance status or lack of appropriate donors. We identified 26 (5%) patients who received HSCT in our cohort. However, patients in need of HSCTs at PHD are sent to either to HDSF or Helsinki for the procedure. As we have no data from Helsinki, we may be missing some HSCT patients from PHD. On average, HSCT receiving patients were younger and had less comorbidities (Table 1).

The treatment options for MDS patients unfit for HSCT are limited to supportive care (e.g., blood transfusions, EPO), the hypomethylating agent azacitidine, and lenalidomide for patients with chromosome 5q deletion. Azacitidine was given to 95 (22%) patients in our study, of which 19 were given azacitidine in association with or after HSCT. All patients who received azacitidine before HSCT received it within a year before the transplant and most received it as an induction, i.e., 6 cycles pre transplant. 72 (17%) patients then received azacitidine independently of HSCT (Table 2). HSCT group was youngest with median age 63.3 years (IQR: 60.9-65.5). Azacitidine receiving group was older at median index 72.6 years (IQR: 66.6-77.4) and median initiation of Azacitidine at 73.1 years (IQR: 67.2-77.8).

**Table 2.** Treatment choices.

| Treatment | Treated at SHC |  | Azacitidine treated |  |  |  | HSCT treated |  |  |  |
| --- | --- | --- | --- | --- | --- | --- | --- | --- | --- | --- |
|  |  |  | complete follow-up |  | during/ after treatment |  | complete follow-up |  | during/ after treatment |  |
|  | n | % | n | % | n | % | n | % | n | % |
| **Allo-HSCT** | 26 | 6,1 | 0 | 0 | 0 | 0 | 26 | 100 | 26 | 100 |
| **AZA** | 91 | 21,5 | 72 | 100 | 72 | 100 | 19 | 73,1 | 12 | 46,2 |
| **BT** | 380 | 89,6 | 63 | 87,5 | 62 | 86,1 | 26 | 100 | 24 | 92,3 |
| **EPO** | 133 | 31,4 | 14 | 19,4 | 8 | 11,1 | 7 | 26,9 | 7 | 26,9 |
| **LEN** | 8 | 1,9 | < 5 | - | < 5 | - | < 5 | - | < 5 | - |

'Treated at SHC' = patients receiving active treatment and specialised healthcare, 'complete follow-up' = time from index until end of follow-up i.e., death, AML transformation or end of study (31.12.2019), 'during/after treatment' = time between azacitidine treatment initiation and end of follow-up, AZA=azacitidine, BT=blood transfusion, EPO=erythropoietin, LEN=lenalidomide.

Red blood cell transfusions (RBCT) were given to 380 (67%) patients. MDS patients often develop dependency on blood transfusions (BT), and indeed, patients received 7 units per patient year (PPY), which increased to 16 units PPY for patients receiving azacitidine, posing a significant burden to both patients and healthcare. Median minimum hemoglobin value within one week pre first RBCT was 82 g/l (IQR: 76-90). Platelets were given to 123 patients (22%), with a frequency of 2.6 units PPY (Table 3).

**Table 3.** Blood transfusions.

| Product type | Subgroup |  | Patients transfused (n) | Days with tranfusion PPY | Units transfused PPY |
| --- | --- | --- | --- | --- | --- |
| Red blood cell | All patients | Allo-HSCT | 26 | 6.0 | 10.0 |
|  |  | Treated at SHC | 380 | 6.4 | 9.1 |
|  | Azacitidine treated patients | AZA any time | 63 | 10.8 | 15.4 |
|  |  | During AZA | 57 | 11.4 | 16.3 |
|  |  | Post-AZA | 46 | 12.1 | 16.8 |
|  |  | Pre-AZA | 26 | 8.4 | 12.7 |
| Platelet | All patients | Allo-HSCT | 23 | 4.1 | 7.7 |
|  |  | Treated at SHC | 123 | 1.4 | 2.6 |
|  | Azacitidine treated patients | AZA any time | 34 | 2.3 | 3.8 |
|  |  | During AZA | 21 | 2.2 | 3.7 |
|  |  | Post-AZA | 23 | 3.7 | 6.0 |
|  |  | Pre-AZA | 5 | 0.9 | 1.4 |

Days with transfusion PPY = number of distinct days a patient received blood transfusion(s) per patient year, units transfused PPY = number of blood units per patient year, AZA any time = Azacitidine receiving patients during whole follow-up, During AZA = during azacitidine treatment, Post-AZA = after azacitidine treatment, Pre-AZA= before azacitidine treatment.

### Outcomes

Median survival of all MDS patients was 26.9 months (95% CI: 23.6-35.1) (Figure 1A). Patients whose disease required treatment at specialised care had a poorer survival than those treated with watchful observation or supportive care at primary care setting (Figure 1B). Median survival for patients treated at specialised care was 22.5 months (95% CI: 17.8-24.8), whereas for patients who are treated at primary care / not treated, have a median survival of 79.7 months (95% CI: 49.7 – not reached), supporting the notion that these patients are indeed low/ medium risk patients. Survival of patients who undergo HSCT is significantly improved, with 73% of patients alive at 24 months (95% CI: 49-87; median survival not reached during study period). As expected, overall survival was also strongly dependent on age at index, with 86,5% (95% CI:76,96-97,21) of under 60-year-olds alive at 24 months, whereas only 43,1% (95% CI: 36,48-50,87) of over 80-year-olds were alive at 24 months (Figure 1C).

**Figure 1.**
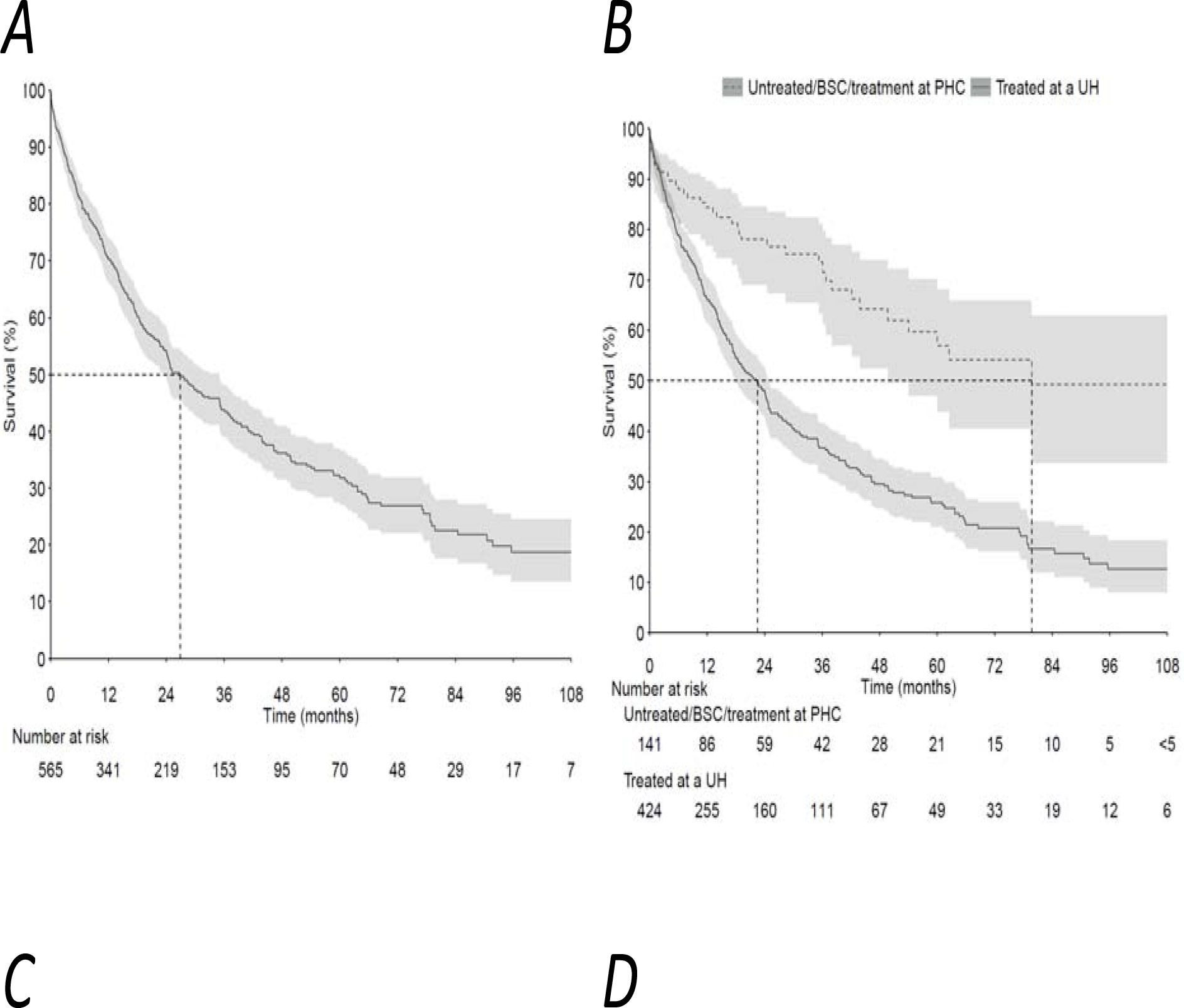
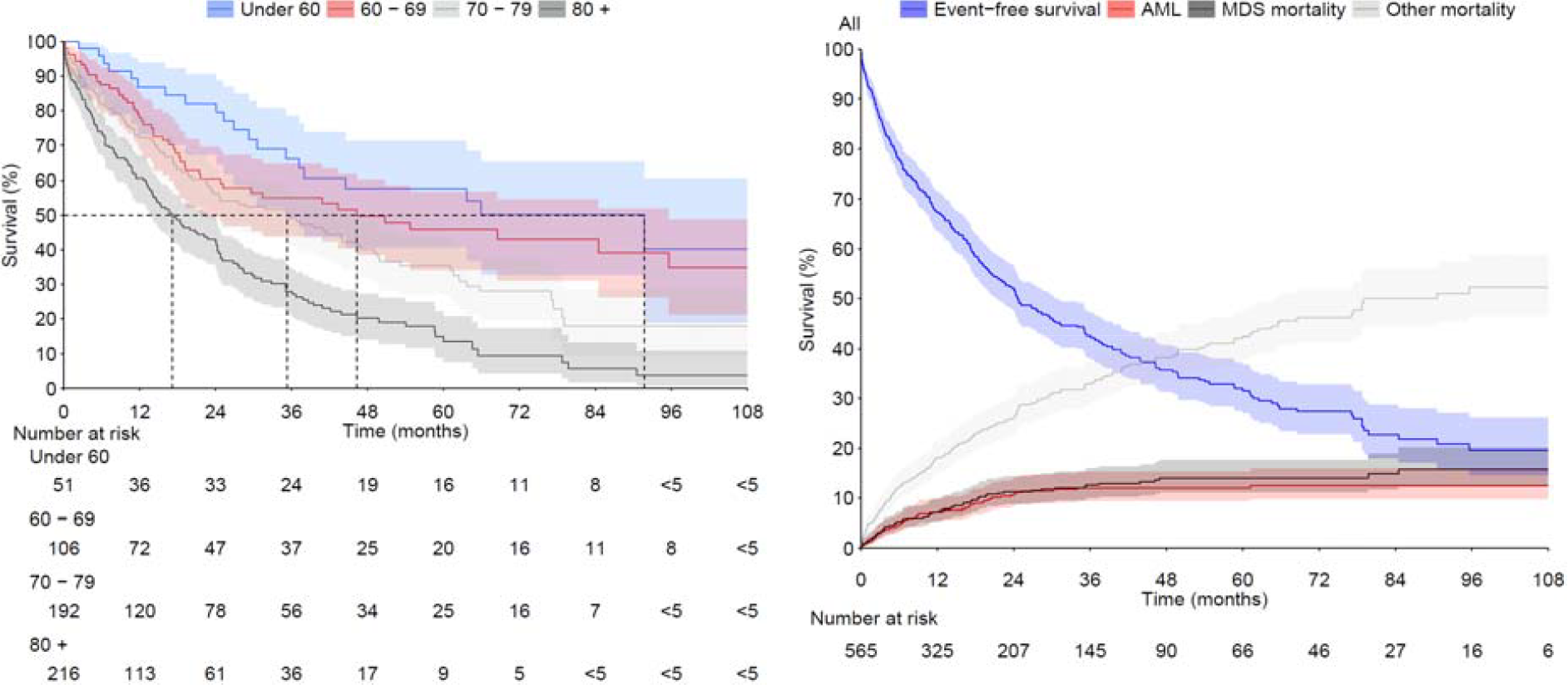
**A)** OS of all MDS patients, both specialised care treated and those who were treated in primary care. Dashed line represents median survival **B)** OS of MDS patients who received treatment at specialty healthcare (dashed line) and who did not (solid line). **C)** Overall survival of MDS patients in different age groups. **D)** Competing risk model of all MDS patients to AML transformation (red line), MDS caused mortality (black line) and other cause mortality (grey line). Blue line represents proportion of live, not-AML-transformed patients.

A major risk for MDS patients is progression into AML. 60 MDS patients (10%) in our study progressed to AML (Figure 1D). These patients had a poor outcome, with median OS of just 3.9 months (95% CI: 2.1-5.3). MDS was the underlying or immediate cause of death for 17% of the cohort. The large proportion of other (non-MDS) causes of death is indicative of the fragile nature of these patients (Figure 1D). For the “other mortality” group the most common causes of death were pneumonia (29 patients, 13% of other mortality), chronic ischemic heart disease (24 patients, 11% of other mortality), heart failure (13 patients, 6% of other mortality) and myocardial infarction (11 patients, 5% of other mortality). Of note, 25 patients who did not have a record of AML diagnosis during their lifetime, had AML recorded as a cause of death.

For the subgroup initiating azacitidine treatment median OS is 22.6 months (95% CI: 17.3-25.2) (Figure 2A), close to that of all specialised care treated patients (Figure 1B). However, azacitidine treated patients had a higher risk of developing AML, and proportionally higher risk of dying of MDS (Figure 2B).

**Figure 2.**
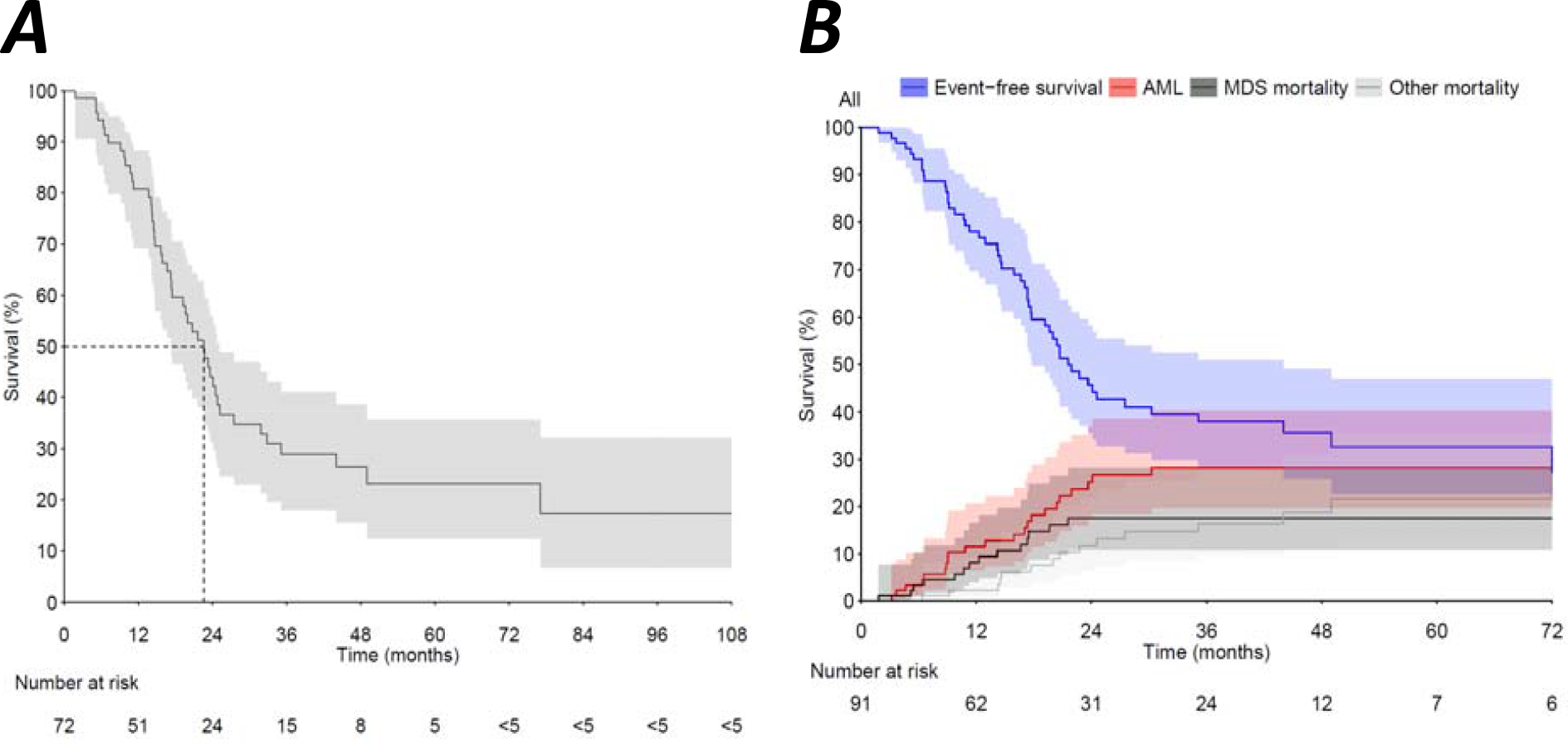
**A)** OS for azacitidine treated patients from the beginning of azacitidine treatment. **B)** Time to event analysis of azacitidine treated patients from the beginning of azacitidine treatment. AML= transformation to acute myeloid leukemia, MDS-mortality = MDS specific mortality where MDS is an immediate or underlying cause of death, Other mortality = cause of death other than MDS.

### HCRU

The cost of care for MDS patients in Finland is substantial. Specialised healthcare resource utilization of MDS patients was calculated per patient year and divided to inpatient and outpatient HCRU, as well as MDS-specific (visits where MDS is recorded as diagnosis) and other cause costs. The economic burden of MDS treatment consists of both hospitalizations and outpatient visits, and is higher for azacitidine treated patients (Figures 3 and 4). On average, SHC treated MDS patients spent 42 days (95% CI: 37-48 days) in hospital per year (inpatient and outpatient days combined), whereas for azacitidine treated patients the number of days in hospital per year is 87 (95% CI: 73-103 days). For all SHC treated MDS patients approximately one third of total per patient year costs were MDS specific (total cost PPY 17 563€, 95% CI: 15 488-19 898€, MDS specific costs 4 704€, 95% CI: 3 747-5 846€), whereas for azacitidine treated patients the costs were more strongly driven by MDS, with approximately half of costs being MDS specific (total costs for MDS patients per patient year 33 094€, 95% CI: 27 592-39 875, MDS specific costs 18 824€ 95% CI: 14 060-24 424€).

**Figure 3.**
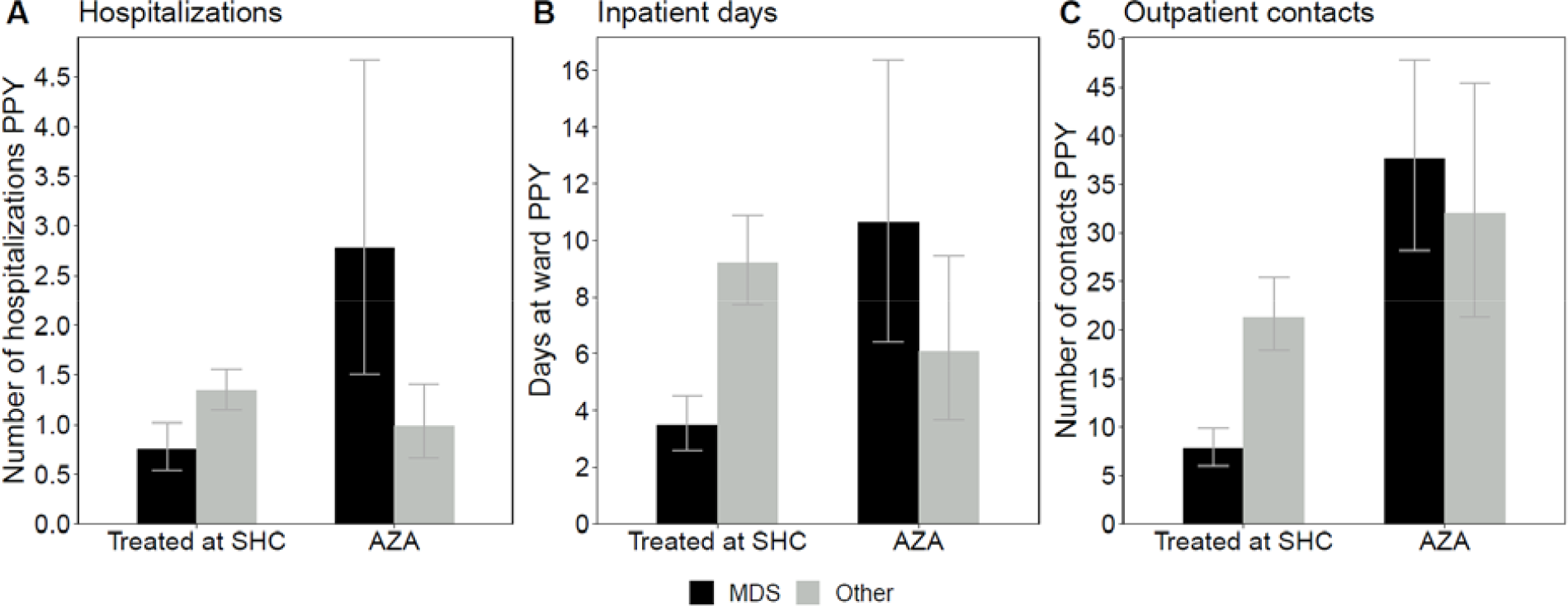
Hospitalizations (panel A), inpatient days (i.e. days at ward; panel B) and outpatient contacts (panel C) per patient year according MDS specific and other health care use in all MDS patients treated at specialty health care and AZA treated MDS patients.

**Figure 4.**
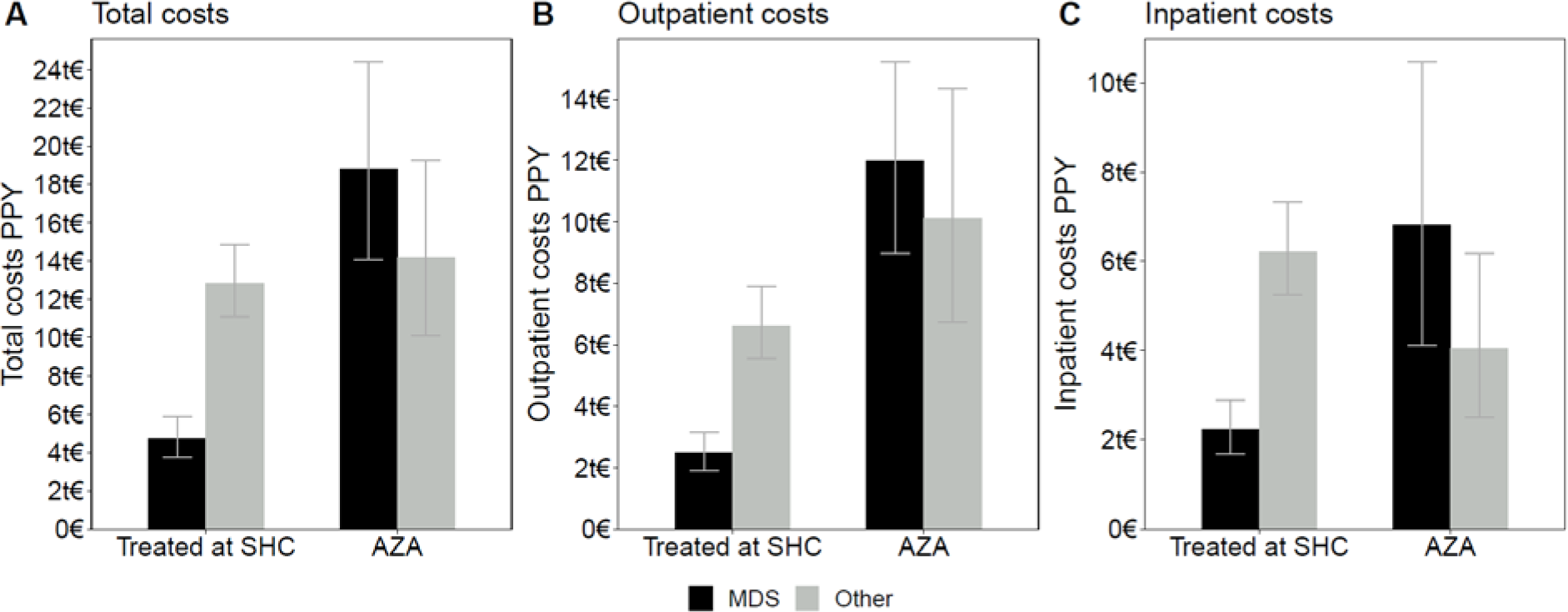
HCRU related MDS/CMML specific costs and other costs per patient year in all MDS patients treated at specialty health care and in AZA treated MDS patients. Total costs are reported in panel A while the costs originating from hospitalizations and outpatient contacts are reported separately in panels B and C, respectively.

## Discussion

Diagnosis of MDS is increasingly attempted early in the course of the disease for all patients, also for asymptomatic and fragile patients. This has led to changes in the patient landscape of MDS and a need to describe these patients, their treatment, and outcomes. Most mild or asymptomatic patients in early stages of MDS are initially treated with watchful observation or limited supportive care but may be treated more aggressively if / when disease progresses. Patients with low life expectancy or high comorbidity burden who do not tolerate high intensity treatment receive supportive / palliative care only. Fit, high-risk and/or symptomatic patients may be treated with azacitidine, and the youngest most fit are considered for HSCT, the only possibly curative treatment. The decision to include a patient in any of these groups should be evidence based, lead to best possible outcomes for the patient, as well as be based on justifiable usage of healthcare resources.

Here, we describe MDS patients with different treatment options in two hospital districts in Finland. We show that the majority of the patients are treated with blood transfusions and younger, fitter patients with allo-HSCT and / or azacitidine. The decision of treatment option is reflected in the outcomes for the different treatment groups, where those treated with watchful observation have a median OS of more than 6 years, and those receiving specialised care less than 2 years (Figure 1). No major differences in characteristics (e.g., age, comorbidities) were seen between the patient groups treated at specialised care and those who were not (Table 1), indicating that the decision to treat was based on disease characteristics. Azacitidine treated patients have a similar median OS of less than 2 years but have a higher probability of their disease progressing to AML and dying of MDS (Figure 2). Importantly, the treatment decision is also reflected in the costs of treatment (Figure 3). Patients chosen for higher intensity treatment need to therefore tolerate the treatments, blood counts need to improve, and the appropriateness of treatment needs to be regularly assessed. Our data concurs well with other previously published real world studies with MDS (3,10,11).

This study was based on data from electronic health records from two Finnish hospital district data lakes and Statistics Finland, accessed via the national health and social data authority Findata. This is a fast and affordable method of study as the data is extracted automatically and is mostly in structured format, ready for analysis with minimal data cleaning steps. However, for more non-structurally available variables, such as IPSS-R risk score, a traditional chart review would still be required. To our knowledge this is the first report of real-world data on MDS in Finland.

This study included data on only specialised healthcare. Watchful observation patients are often treated at primary care, and during the study period also some blood transfusions may have taken place at primary care, we may miss some of their treatment and costs. This study is also limited by its real-world nature such as coding practices in clinical care and is subject to missing data. The nature of automated data access also limits the scope of data, so that here e.g. IPSS scores are missing.

The choice of medication for MDS patients remains limited, and therefore poses a significant therapeutic challenge. While the treatment of haematological cancers has improved significantly during the past decade, the drug development for MDS lags behind, and new alternatives are needed.

## Data Availability

The data that support the findings of this study were collected by following the guidance and application process of Findata, the central permission authority in Finland. Restrictions apply to the data, where only persons named in the study permission have access to the pseudonymized micro data for the current study, and so are not publicly available.

## Acknowledgements

The authors would like to acknowledge Riikka Mattila, PhD and Scientific Advisor at Medaffcon Oy for editorial- and writing assistance, which was funded by Takeda Pharma Oy.

## References

1. Jain AG, Elmariah H. BMT for Myelodysplastic Syndrome: When and Where and How. Frontiers in Oncology [Internet]. 2022 [cited 2022 May 11];11. Available from: https://www.frontiersin.org/article/10.3389/fonc.2021.771614

2. Tsutsué S, Suzuki T, Kim H, Crawford B. Real-world assessment of myelodysplastic syndrome: Japanese claims data analysis. Future Oncology. 2022 Jan;18(1):93–104.

3. Rozema J, Hoogendoorn M, Kibbelaar R, van den Berg E, Veeger N, van Roon E, et al. Comorbidities and malignancies negatively affect survival in myelodysplastic syndromes: a population-based study. Blood Advances. 2021 Mar 3;5(5):1344–51.

4. Joshi N, Kale H, Corman S, Wert T, Hill K, Zeidan AM. Direct Medical Costs Associated With Treatment Nonpersistence in Patients With Higher-Risk Myelodysplastic Syndromes Receiving Hypomethylating Agents: A Large Retrospective Cohort Analysis. Clinical Lymphoma, Myeloma and Leukemia. 2021 Mar 1;21(3):e248–54.

5. Quan H, Sundararajan V, Halfon P, Fong A, Burnand B, Luthi JC, et al. Coding Algorithms for Defining Comorbidities in ICD-9-CM and ICD-10 Administrative Data. Medical Care. 2005 Nov;43(11):1130–9.

6. Mäklin S. Terveyden-ja sosiaalihuollon yksikkökustannukset Suomessa vuonna 2017. 2017;55.

7. Therneau T. A package for survival analysis in R. :97.

8. Thygesen SK, Christiansen CF, Christensen S, Lash TL, Sørensen HT. The predictive value of ICD-10 diagnostic coding used to assess Charlson comorbidity index conditions in the population-based Danish National Registry of Patients. BMC Med Res Methodol. 2011 Dec;11(1):83.

9. PDQ Adult Treatment Editorial Board. Myelodysplastic Syndromes Treatment (PDQ®): Health Professional Version. In: PDQ Cancer Information Summaries [Internet]. Bethesda (MD): National Cancer Institute (US); 2002 [cited 2022 May 16]. Available from: http://www.ncbi.nlm.nih.gov/books/NBK66015/

10. Balleari E, Salvetti C, Del Corso L, Filiberti R, Bacigalupo A, Bellodi A, et al. Age and comorbidities deeply impact on clinical outcome of patients with myelodysplastic syndromes. Leukemia Research. 2015 Aug 1;39(8):846–52.

11. Tsutsué S, Suzuki T, Kim H, Crawford B. Real-world assessment of myelodysplastic syndrome: Japanese claims data analysis. Future Oncology [Internet]. 2021 Oct 15 [cited 2022 Mar 28]; Available from: https://www.futuremedicine.com/doi/full/10.2217/fon-2021-0988

